# Left Ventricular Reverse Remodeling After Revascularization and Its Predictive Role for Survival

**DOI:** 10.1101/2023.10.31.23297881

**Authors:** Shaoping Wang, Bijan J. Borah, Shujuan Cheng, Shiying Li, Yanci Liu, Xiaoyan Gu, Jinhua Li, Yi Lyu, Jinghua Liu

**Affiliations:** Department of Cardiology, Beijing Anzhen Hospital, Capital Medical University, Beijing Institute of Heart Lung and Blood Vessel Diseases, Beijing, China; Department of Health Sciences Research, Mayo Clinic, Rochester, Minnesota, USA; Robert D. and Patricia E. Kern Center for Science of Health Care Delivery, Mayo Clinic, Rochester, Minnesota, USA; Department of Echocardiography, Beijing Anzhen Hospital, Capital Medical University, Beijing Institute of Heart Lung and Blood Vessel Diseases, Beijing, China; Department of Cardiovascular Surgery, Beijing Anzhen Hospital, Capital Medical University, Beijing Institute of Heart Lung and Blood Vessel Diseases, Beijing, China; Department of Anesthesiology, Minhang Hospital, Fudan University, Shanghai, China

**Author notes:** Corresponding Author: <u>Jinghua Liu, MD, PhD</u>, Department of Cardiology, Beijing Anzhen Hospital, Capital Medical University, No 2 Anzhen Road, Chaoyang District, Beijing, 100029 China. <u>Yi Lyu MD</u>, Department of Anesthesiology, Minhang Hospital, Fudan University, No 180 Xinsong Road, Minhang District, Shanghai, 201199.

**Keywords:** bypass, ejection fraction, heart failure, revascularization, stents

## Abstract

**Aims:** For patients with ischemic heart failure who underwent revascularization, ejection fraction (EF) improvement is a major predictor of survival benefit. However, the association between left ventricular (LV) remodeling and outcomes has not been well-established. The aim of the study is to investigate the extent of LV remodeling after revascularization and its predictive role for long-term survival.

**Methods:** Patients with reduced EF (≤40%), who underwent either coronary artery bypass grafting or percutaneous coronary intervention, and had echocardiography reassessment 3 months after revascularization were enrolled in a real-world cohort study (No. ChiCTR2100044378). Patients were categorized into 4 groups according to whether LV end-systolic dimension (LVESD) reduction was ≤7% or >7%, and absolute EF improvement ≤5% or >5%

**Results:** A total of 923 patients were identified. The percentage of LVESD reduction was 4.5±18.4%. The median follow-up time was 3.4 years, during which 123 patients died. Patients with greater percentage of LVESD reduction had lower risk of all-cause death (hazard ratio [HR] per 1% decrement in LVESD, 0.98; 95% CI, 0.97-0.99; *P*<.001). A reduction in LVESD of 7.2% was the optimal cutoff value to predict survival. Compared to patients with LVESD reduced and EF improved, 2.11-fold (95% CI, 1.04-4.29), 3.56-fold (95% CI, 1.60-7.91), and 7.54-fold (95% CI, 4.20-13.53) higher mortality were found in LVESD unreduced but EF improved, LVESD reduced but EF unimproved, and LVESD unreduced and EF unimproved group, respectively.

**Conclusions:** After revascularization among patients with ischemic HF, a reduction in LVESD of 7% signifies clinically relevant revers remodeling. Combination of EF improvement and LVESD reduction might be more clinically precise approach of risk stratification in this population.

**Clinical Trial Registration:** The name of the registry: Coronary Revascularization in Patients with Ischemic Heart Failure and Prevention of Sudden Cardiac Death.

Registration number: ChiCTR2100044378 (http://www.chictr.org.cn).

## Introduction

Coronary artery disease (CAD) is the most common cause of heart failure (HF) and is associated with substantial morbidity, mortality, and expenditure of health care resources in world wild.^1–3^ Revascularization by either coronary artery bypass grafting (CABG) or percutaneous coronary intervention (PCI) may attenuate myocardial ischemia and reverse left ventricular (LV) adverse remodeling, thus improve long-term outcomes of patients with LV dysfunction in comparison to optimal medical therapy alone. ^4–7^ However, not all patients with ischemic HF had LV reversal remodeling and outcomes improvement after revascularization.^8–10^ Echocardiography reassessment 3 months after revascularization is recommended to evaluate the effect of revascularization and risk stratify for patient management.^11, 12^ Improvement in LV ejection fraction (EF) has been reported to be a major predictor of survival benefit in patients with HF and reduced EF, irrespective of such an improvement in EF is facilitated by revascularization, device therapy, or medical therapy alone.^8, 13–17^ In our previous study, patients with EF improvement as defined by absolute increase in EF >5% after revascularization were associated with over 50% reduction of risk of all-cause death in comparison to those without EF improvement as defined by absolute increase in EF ≤5%.^8^

Besides change of LV systolic function, change of left ventricular size is another important index of LV remodeling. Reversed LV remodeling by drug or device therapy has been reported to be associated with improved long-term survival of patients with HF and reduced EF.^15, 18^ A decrement of 10 ml in the mean LV end-systolic volume (LVESV) corresponded to a relative odds ratio of 0.96 for mortality in a meta-analysis.^15^ In this meta-analysis, patients who received either drug or device therapies in randomized controlled trials were included together for remodeling and mortality analyzing. However, response of LV remodeling might be different between drug and device therapy. The magnitude of LV reverse remodeling after cardiac resynchronization therapy (CRT) was reported to be a >20% reduction in LVESV, which is much larger than that observed in medical therapy.^14, 19, 20^ So far, to our knowledge, for patient with ischemic HF who underwent revascularization, the extent of LV remodeling after revascularization and its predictive role in outcomes has not been well-established.

Therefore, this study was performed to investigate 1) the change of LV dimension following revascularization among patients with CAD and preoperative EF ≤40%; 2) the association between LV dimension reduction and EF improvement; 3) what extent of LV reverse remodeling predicts improved outcome in this population.

## Methods

### Patient Selection and Definition

This was a real-world cohort study and registered in Chinese Clinical Trial Registry (No. ChiCTR2100044378). The study protocol was approved by the ethics committee of Beijing Anzhen Hospital. Patients who underwent CABG or PCI with drug-eluting stent because of CAD in Beijing Anzhen Hospital from January 1, 2005, to December 31, 2014 were screened. Patients were enrolled if they had initial reduced EF (≤40%), and underwent isolated CABG or PCI, and had repeated echocardiographic measurements during follow-up. Patients were excluded if they were diagnosed as ST-segment elevation myocardial infarction (MI), died within 3 months after revascularization, and had only one record of echocardiographic reassessment within 3 months after CABG or PCI.

According to the absolute change in EF between preoperative and postoperative measurement, patients were categorized: 1) EF unimproved group (absolute increase in EF ≤5%); 2) EF improved group (absolute increase in EF >5%).^8, 13^ The percentage of LVESD reduction was defined as below: LVESD reduction, % = (postoperative LVESD – preoperative LVESD) / preoperative LVESD*100. According to the percentage of LVESD reduction, patients were categorized: 1) LVESD unreduced group (absolute decrease in percentage of LVESD reduction ≤7%); 2) LVESD reduced group (absolute decrease in percentage of LVESD >7%).

### Data Collection and Definitions

The clinical, laboratory, echocardiographic data and medical therapy were recorded from hospital medical records. Baseline echocardiographic data was captured within 30 days before PCI or CABG. Follow-up echocardiographic data were defined as the first measurement 3 months^11, 12, 21^ after revascularization assessed in Beijing Anzhen Hospital. Complete revascularization was defined as successful PCI (residual stenosis of <30%) of all angiographically significant lesions (≥70% diameter stenosis) in 3 coronary arteries and their major branches. For CABG, grafting of every primary coronary artery with ≥70% diameter stenosis was accepted as complete revascularization.

Outcome data were obtained from medical records at Beijing Anzhen Hospital and through telephone follow up. Death was regarded as cardiovascular in origin unless obvious non-cardiovascular causes could be identified. Any death during hospitalization for repeat coronary revascularization was regarded as cardiovascular death. The follow-up time for patients started at the time of the first available EF measurement^13, 22, 23^.

### Statistical Analysis

Continuous variables were presented as mean ± standard deviation (SD) or median (interquartile range). The categorical variables were reported as counts and percentages. Baseline characteristic and echocardiography parameters were compared between LV end-systolic diameter (LVESD) reduced and LVESD unreduced group by using a student *t* test, rank sum test, χ^2^ test, or Fisher’s exact test as appropriate. Receiver-operating-characteristic (ROC) curve was analyzed to assess the best cutoff value of LVESD reduction, LV end-diastolic diameter (LVEDD) reduction and EF improvement to predict all-cause death. Cumulative incidences were estimated with Kaplan-Meier method and compared using the log-rank test. The risks of outcomes were analyzed using a Cox proportional hazards regression model. The model was adjusted with age, sex, hypertension, diabetes, renal function, history of MI, treatment with PCI or CABG, and complete revascularization.

All statistical analyses were based on 2-tailed tests. *P* values less than .05 were considered statistically significant. Statistical analyses were performed with Stata version 14.0 (StataCorp LLC).

## Results

### Baseline Characteristics

Among 1,781 initially identified patients, 78 patients who died within 3 months after revascularization, 780 patients were further excluded because echocardiography was not evaluated 3 months after revascularization. Finally, 923 patients who had an initial EF ≤40% and had echocardiography reassessment 3 months after revascularization were enrolled in this study.

The average age at baseline was 64.7±10.8 years. Men comprised 83.4% of all subjects. 526 (57.0%) received PCI and 397 (43.0%) underwent CABG **(Table 1)**. Mean (SD) preoperative EF was 36.3% (4.3%). EF was changed into 45.2% (11.2%) in average with an absolute 8.9% (11.0%) improvement after revascularization. The average preoperative LVESD was 45.9 (8.2) mm. After revascularization, absolute reduction in LVESD was 2.5 (8.4) mm. Postoperative LVESD thus changed into 43.4 (9.8) mm in average. The percentage of LVESD reduction was 4.5% (18.4%) and the population distribution according to the percentage of LVESD reduction was shown in **Figure 1**. ***Predictive Role of LV Remodeling in Mortality***

**Table 1.**
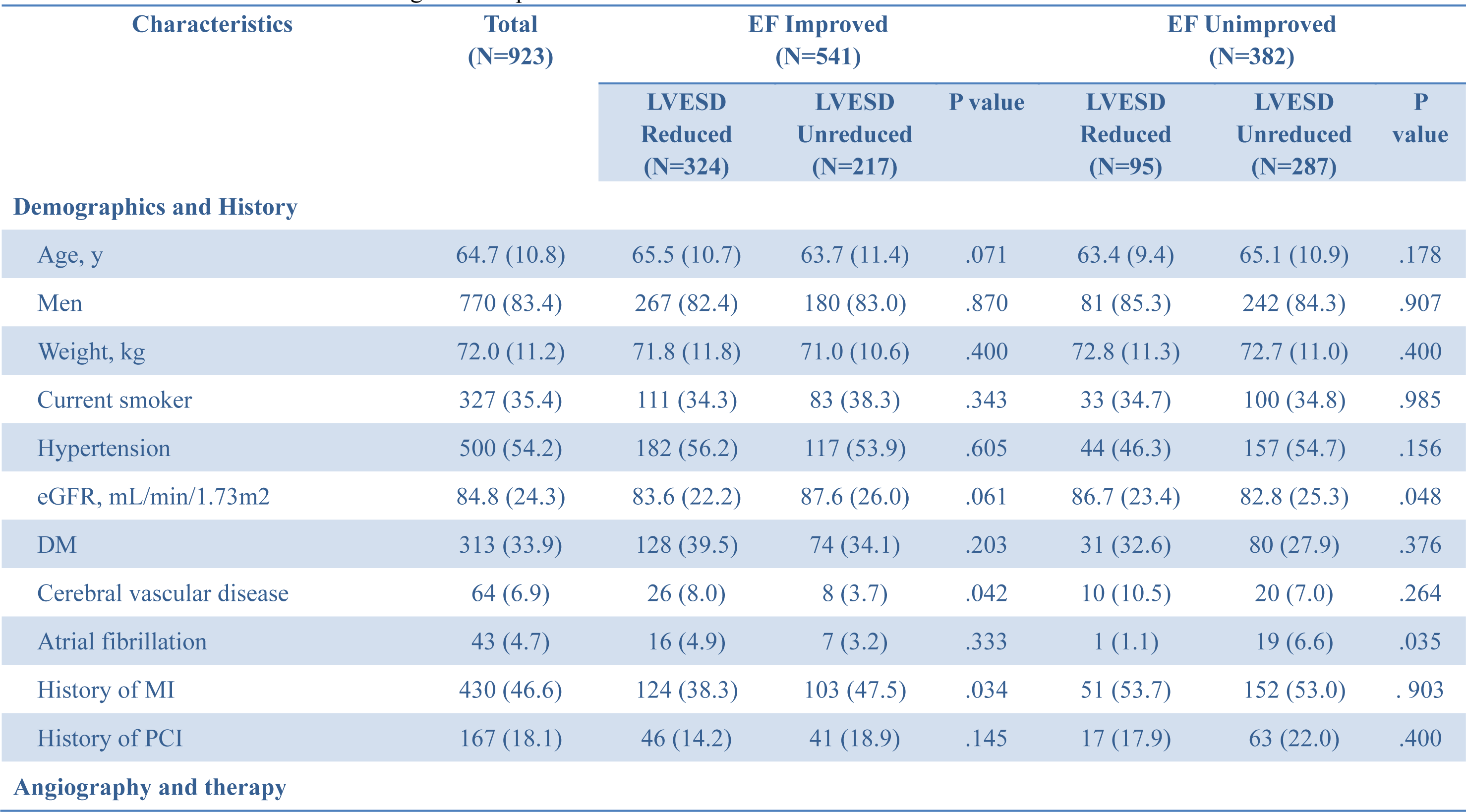
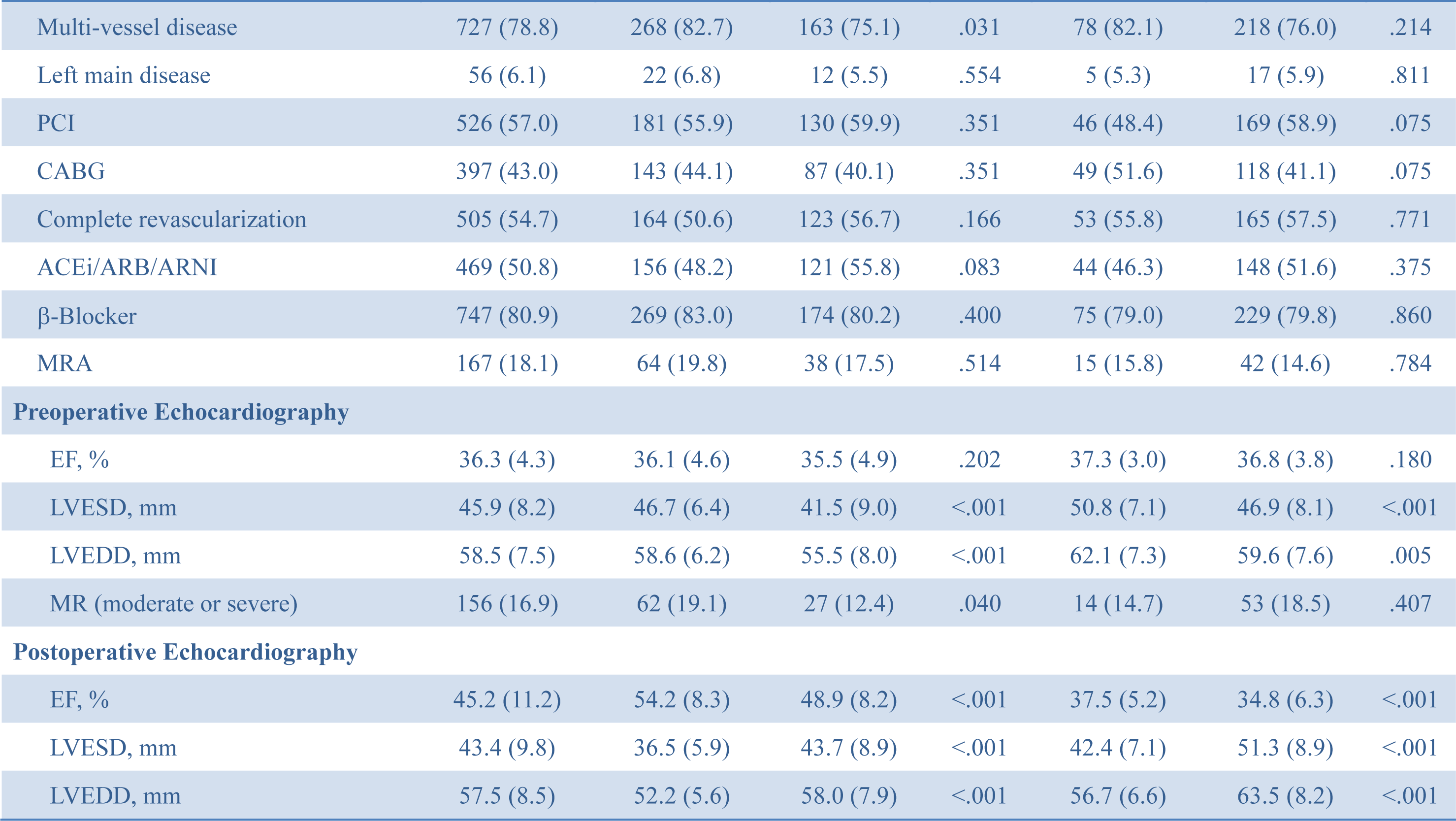
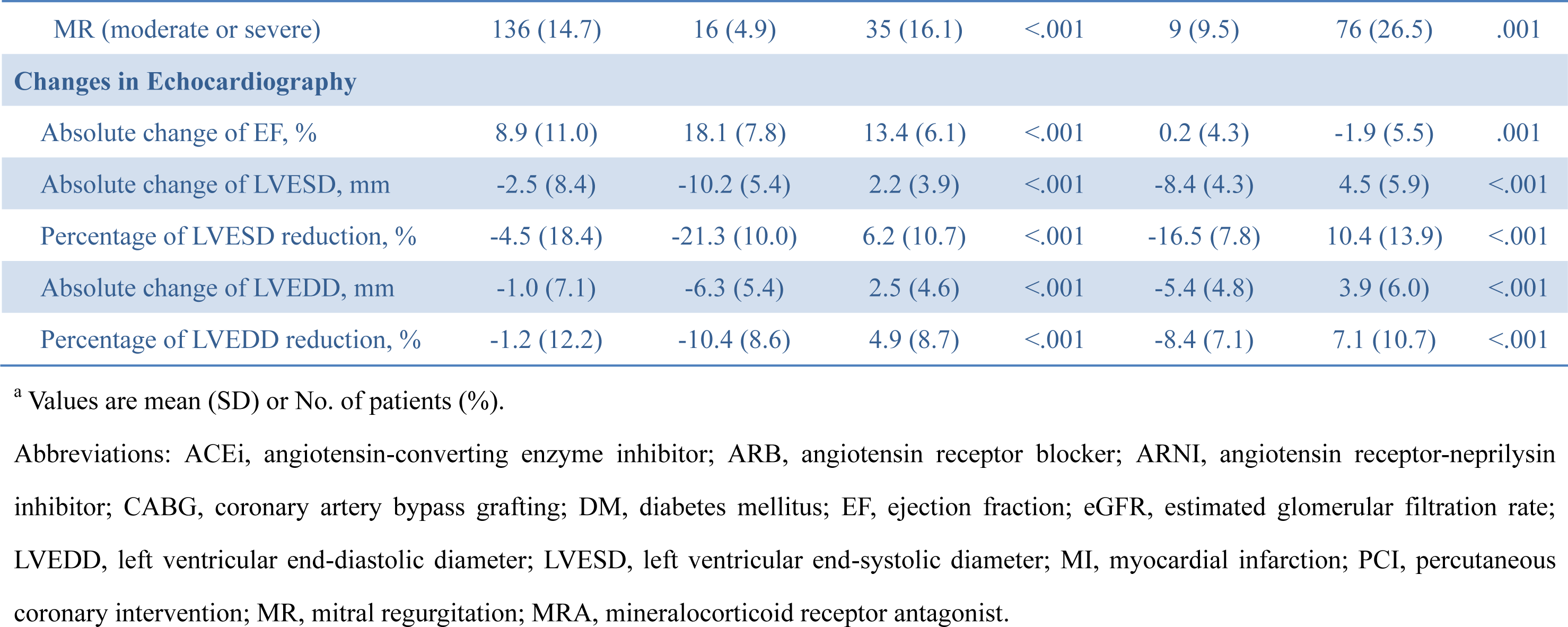
Patient characteristics according to EF improvement and LVESD reduction^a^.

**Figure 1:**
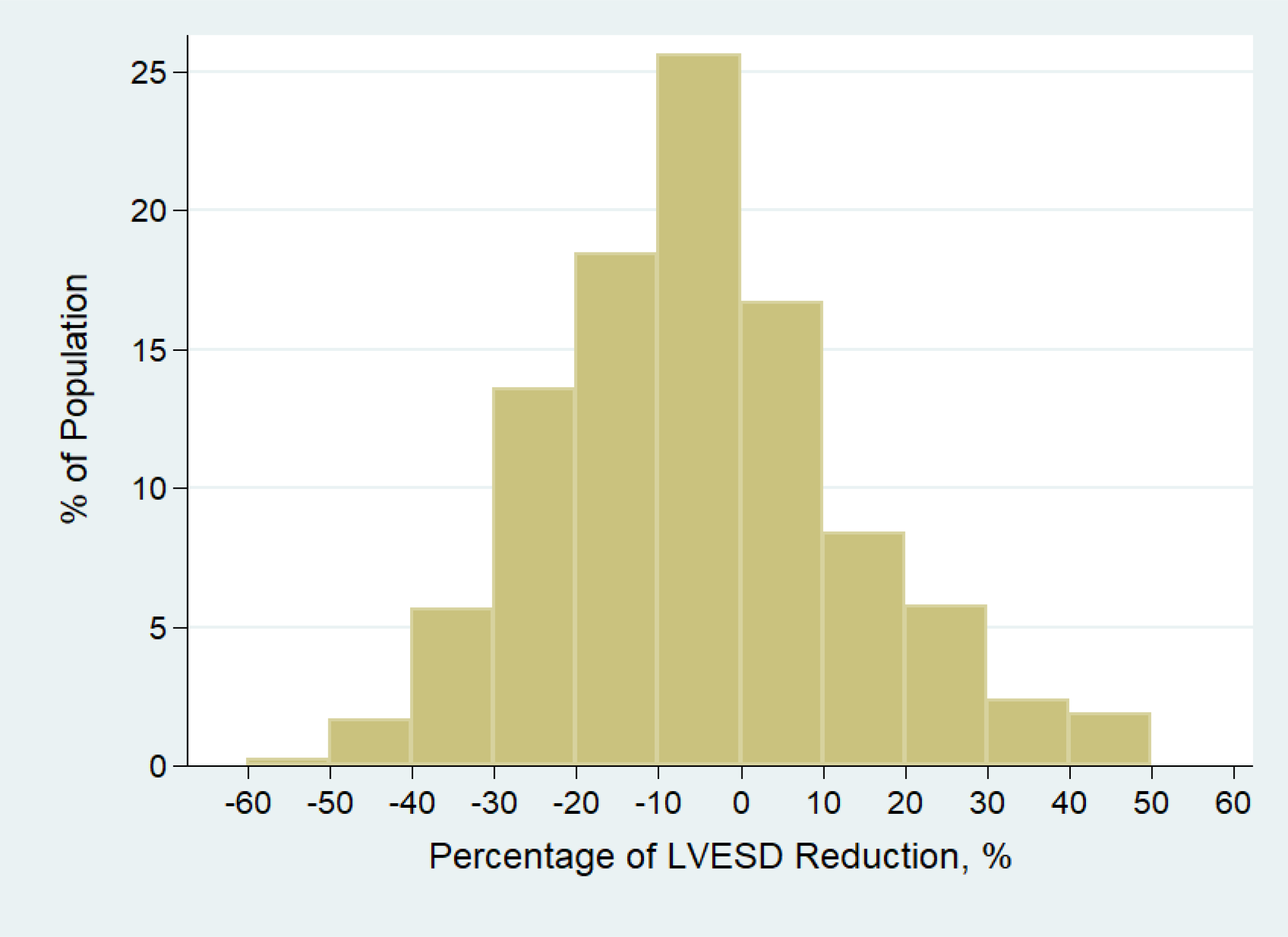
Change of the percentage of LVESD reduction after revascularization. LVESD, left ventricular end-systolic diameter

The median follow-up time was 3.4 years, during which 123 patients died. Of those, 6 (4.9%) died of MI, 40 (32.5%) died of HF, 55 (44.7%) died suddenly, and 22 (17.9%) died of non-cardiac causes.

Patients with greater percentage of LVESD reduction had significant lower risk of all-cause death (hazard ratio [HR] per 1% decrement in LVESD, 0.98; 95% CI, 0.97-0.99; *P*<.001). Based on ROC curve analysis, a reduction in LVESD of 7.2% was identified as the optimal cutoff value to predict long-term survival with area under the curve (AUC) of 0.63 (95% CI, 0.58-0.68) **(Figure 2)**. With this cutoff value, a sensitivity of 71% and a specificity of 48% were obtained to predict all-cause mortality. Unsurprisingly, patients with greater EF improvement had significant lower risk of all-cause death (HR per 1% increment in EF, 0.95; 95% CI, 0.94-0.97; *P*<.001). ROC curve analysis indicated the optimal cutoff value of EF improvement to predict survival was 5.8% with AUC of 0.70 (95% CI, 0.65-0.75) **(Figure 2)**.

**Figure 2:**
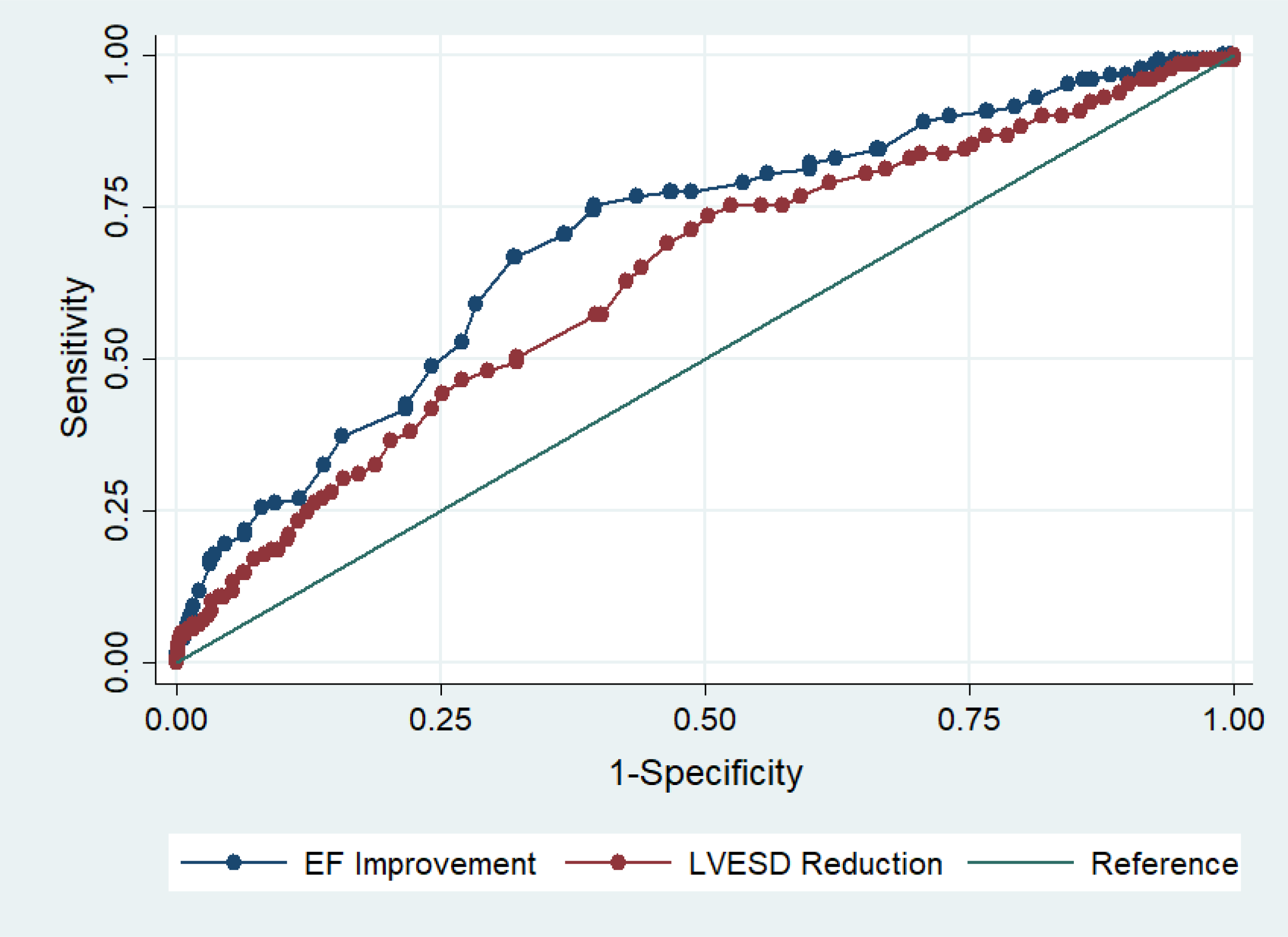
ROC curves for prediction all-cause death. EF, ejection fraction; LVESD, left ventricular end-systolic diameter; ROC, Receiver-operating-characteristic.

419 (45.4%) patients had LVESD reduction > 7% and 504 (54.6%) patients had LVESD reduction ≤7%. Patients with LVESD reduced (LVESD reduction >7%) had lower risk of all-cause death (HR, 0.32; 95% CI, 0.21-0.49; *P*<.001) and cardiovascular mortality (HR, 0.25; 95% CI, 0.15-0.41; *P*<.001) in comparison with patients with LVESD unreduced (LVESD reduction ≤7%) **(Table 2)**. Furthermore, patients with LVESD reduced had significantly lower risk of mortality in EF improved groups (HR, 0.49; 95% CI, 0.24-0.98; *P*=.043) and tended to have significant lower risk in EF unimproved group (HR, 0.56; 95% CI, 0.31-1.01; *P*=.053). In addition, patients with LVESD reduced had significantly lower risk of cardiovascular death both in EF improved and EF unimproved group.

**Table 2.**
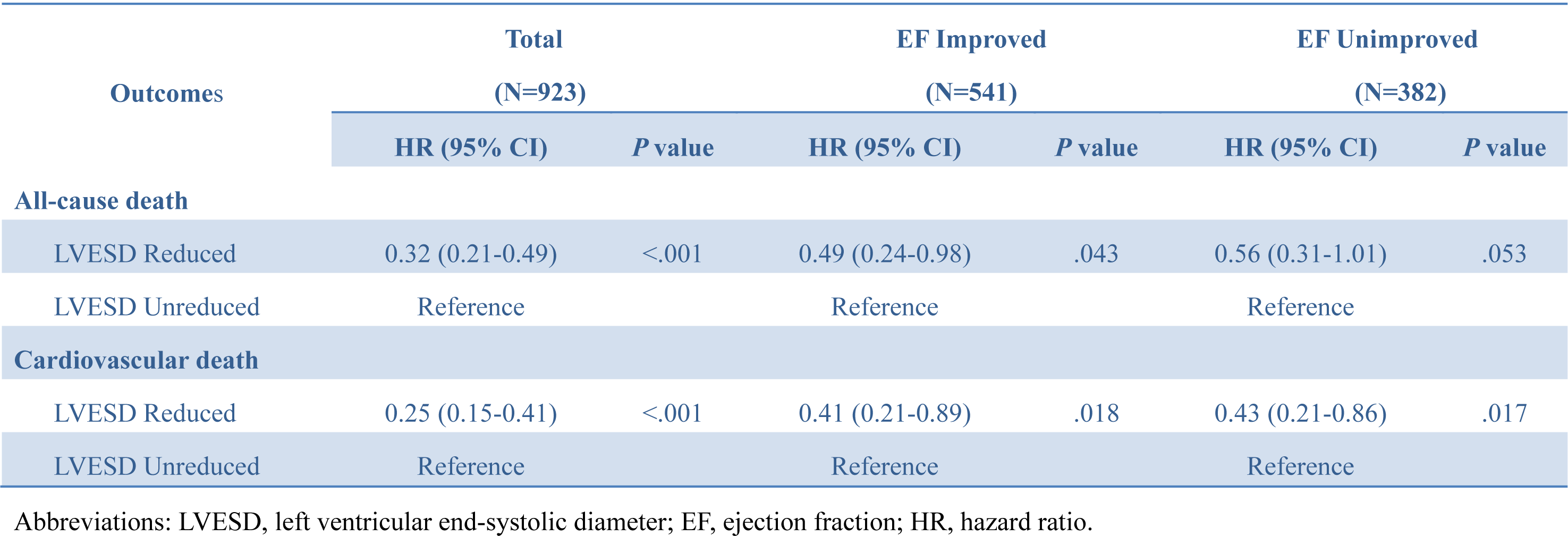
Risk of outcomes (LVESD reduced versus LVESD unreduced)

| Outcomes | Total<br>(N=923) |  | EF Improved<br>(N=541) |  | EF Unimproved<br>(N=382) |  |
| --- | --- | --- | --- | --- | --- | --- |
|  | HR (95% CI) | *P* value | HR (95% CI) | *P* value | HR (95% CI) | *P* value |
| **All-cause death** |  |  |  |  |  |  |
| LVESD Reduced | 0.32 (0.21-0.49) | <.001 | 0.49 (0.24-0.98) | .043 | 0.56 (0.31-1.01) | .053 |
| LVESD Unreduced | Reference |  | Reference |  | Reference |  |
| **Cardiovascular death** |  |  |  |  |  |  |
| LVESD Reduced | 0.25 (0.15-0.41) | <.001 | 0.41 (0.21-0.89) | .018 | 0.43 (0.21-0.86) | .017 |
| LVESD Unreduced | Reference |  | Reference |  | Reference |  |

Abbreviations: LVESD, left ventricular end-systolic diameter; EF, ejection fraction; HR, hazard ratio.

Patients were further categorized into 4 groups according to whether LVESD reduction was ≤7% or >7%, and absolute EF improvement ≤5% or >5% **(Figure 3)**. 324 (35.1%) patients had LVESD reduced and EF improved. 287 (31.1%) had LVESD unreduced and EF unimproved. Patients with LVESD reduced and EF improved had lowest risk of all-cause mortality, whereas patients with LVESD unreduced and EF unimproved had highest risk **(Table 3**, **Figure 4)**. In addition, patients who had LVESD reduced but had EF unimproved tended to have significantly higher risk of mortality in comparison to those who had LVESD unreduced but had EF improved (HR, 1.99; 95% CI, 0.98-4.08; *P*=.058). Those findings persisted in adjusted model.

**Figure 3:**
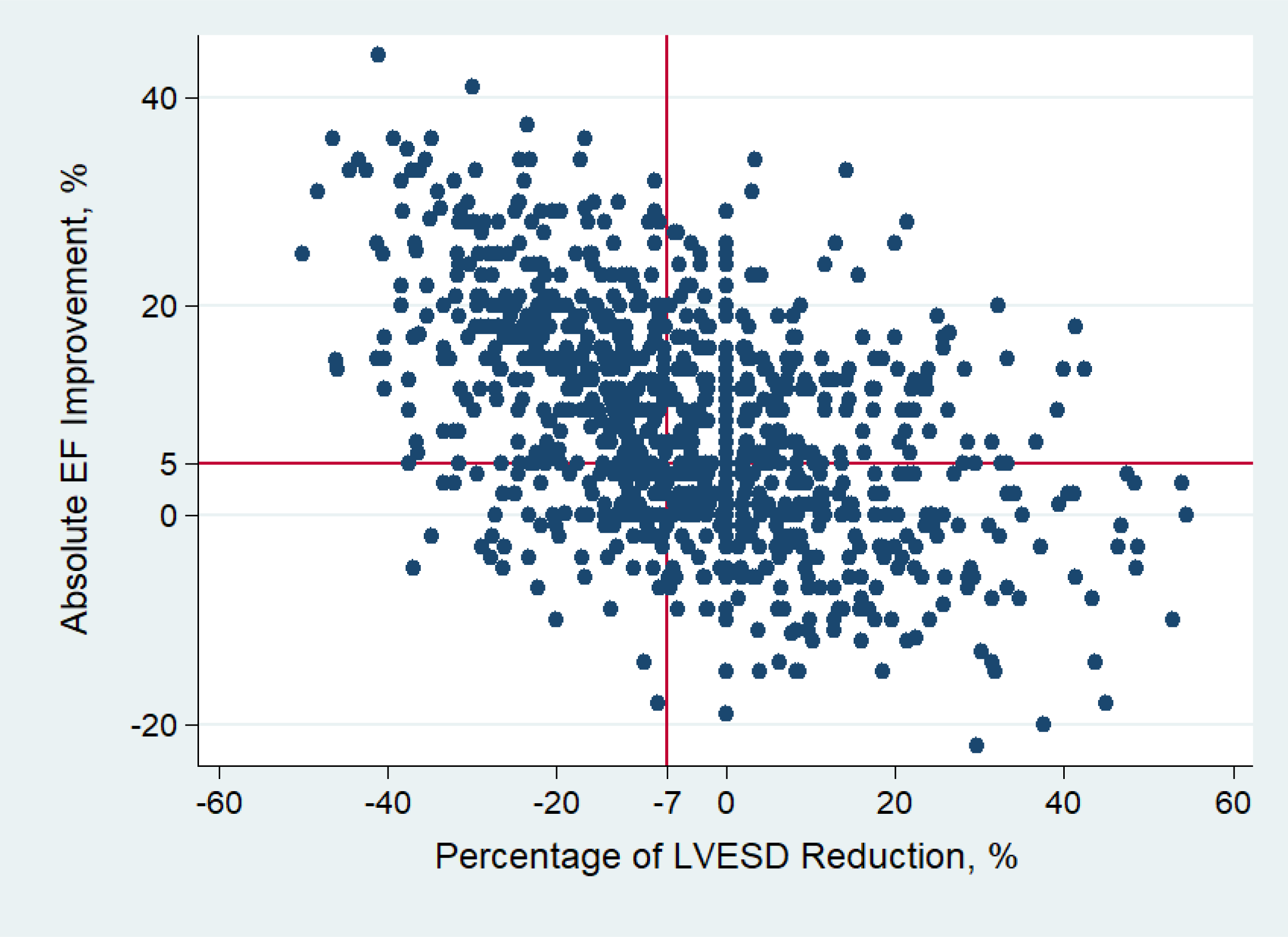
Patient distribution according to the absolute EF improvement and percentage of LVESD reduction after revascularization. EF, ejection fraction; LVESD, left ventricular end-systolic diameter.

**Figure 4:**
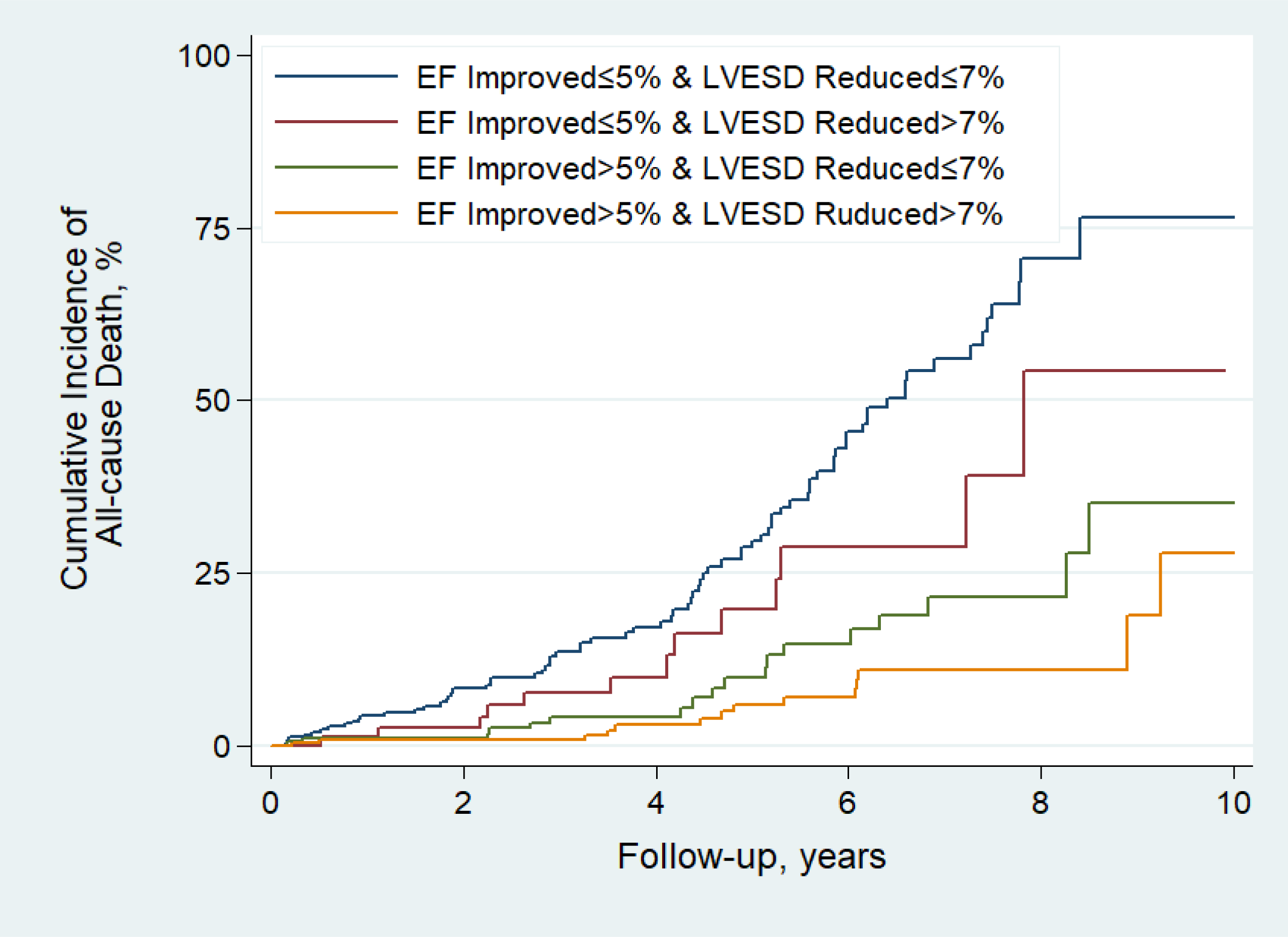
Kaplan-Meier curves estimating incidence of all-cause death. EF, ejection fraction; LVESD, left ventricular end-systolic diameter.

**Table 3.** Risk of all-cause death according to EF improvement and LVESD reduction.

| Outcomes | EF Improved |  | EF Unimproved |  |
| --- | --- | --- | --- | --- |
|  | LVESD Reduced | LVESD Unreduced | LVESD Reduced | LVESD Unreduced |
| Unadjusted | Reference | 2.05 (1.02-4.12) | 4.08 (1.92-8.70) | 7.53 (4.24-13.36) |
| Adjusted <sup>a</sup> | Reference | 2.11 (1.04-4.29) | 3.56 (1.60-7.91) | 7.54 (4.20-13.53) |

Data are HR (95% CI). <sup>a</sup> HRs were adjusted with age, sex, hypertension, diabetes, estimated glomerular filtration rate, history of MI, treatment with PCI or CABG, and complete revascularization.

### Baseline Characteristics according to LV Remodeling

In EF improved group, age at baseline and sex distribution were similar between the LVESD reduced and unreduced groups **(Table 1)**. The LVESD reduced group had a significant lower prevalence of history of MI (38.3% vs 47.5%; P=.034), but higher prevalence of multi-vessel diseases (82.7% vs 75.1%; P=.031). More patients in LVESD reduced group had cerebral vascular disease.

The prevalence of other comorbid conditions was similar. There also was no significant difference in the proportions undergoing revascularization by PCI or CABG, and the groups had similar percentages of complete revascularization. The proportions of medical therapy were similar between the groups. The preoperative EF value was similar between LVESD reduced and unreduced groups **(Table 1)**. The LVESD reduced group had significantly greater LVESD and LVEDD at baseline.

More patients in LVESD reduced group had moderate to severe mitral regurgitation. After revascularization, LVESD reduced group had significantly greater EF improvement and LV size reduction. Thus LVESD reduced group had greater postoperative EF and smaller LVESD and LVEDD finally.

In EF unimproved group, the prevalence of history of MI and multi-vessel disease were similar between LVESD reduced and unreduced group **(Table 1)**. LVESD reduced group had greater value of estimated glomerular filtration rate, and fewer patients in LVESD reduced group had atrial fibrillation. The prevalence of moderate to severe mitral regurgitation at baseline was similar between two groups. Other comparisons of echocardiographic characteristics between two groups were similar to the comparisons in EF improved group.

### Sensitivity Analysis

The clinical implication of LVEDD reduction was further investigated. Patients with greater percentage of LVEDD reduction had significant lower risk of all-cause death (HR per 1% decrement in LVEDD, 0.98; 95% CI, 0.97-1.00; *P*=.008). ROC curve analysis indicated the optimal cutoff value of percentage of LVEDD reduction to predict survival was 1.8% with AUC of 0.55 (95% CI, 0.49-0.60). The AUC of percentage of LVEDD reduction was significantly lower than that of percentage of LVESD reduction (*P*=.008).

## Discussion

This study examined the effect of revascularization on LV reverse remodeling and investigated the clinical significance of LV remodeling in predicting outcomes of patients with CAD and reduced EF. For patients with ischemic HF, revascularization has been recommended to reverse LV adverse remodeling and improve long-term survival.^24^ However, not all patients with ischemic HF could benefit from this treatment strategy. Echocardiography reassessment 3 months after revascularization is necessary to manage patients with precise risk stratification.^11, 12^ Improvement in EF had been reported to be a major predictor of survival benefit in HF patients with both ischemic and non-ischemic etiologies.^8, 13–15^ In current study, among patients with ischemic HF who underwent revascularization, EF improvement was also demonstrated as a strong predictor of long-term survival. Furthermore, current study identified LVESD reduction might be another predictor. Patients with greater percentage of LVESD reduction were associated with lower risk of mortality both in EF improved and EF unimproved groups. Combination of EF improvement and LVESD reduction provided more precise risk stratification. Patients with LVESD reduced and EF improved had lowest risk of all-cause mortality, whereas patients with LVESD unreduced and EF unimproved had worst outcomes.

In current study, the optimal cutoff value of EF improvement to predict survival is of 5.8%, which is close to criteria used in previous publications.^8, 13, 15, 16^ Furthermore, a new cutoff value derived from the ROC curves of mortality prediction concluded that a reduction in LVESD of >7% was clinical relevant. HF is of heterogeneous etiology and the mechanisms of benefit of revascularization may be somewhat different from that of medical therapy and device therapy. The extent of LV reverse remodeling after CRT was much larger than that observed in medical therapy.^14, 19, 20^ For patients who underwent CRT, a 10% reduction in LVESV was identified as the optimal cutoff value to predict long-term survival.^25^ The current study provided evidence that among patients with reduced EF who underwent revascularization, a reduction in LVESD of >7% was clinical significance to assist risk discrimination.

According to whether EF improved or not, and LVESD reduced or not after revascularization, two patient cohorts with inconsistent recovery of LV function and structure were identified. 23.5% patients had EF improved, but had LVESD unreduced. 10.3% patients had EF unimproved, but had LVESD reduced. It is unclear whether this dyssynchronized recovery represents a stage during LV remodeling or it could be the result of underlying molecular events involved in remodeling.^26^ Further studies to continuously examine LV remodeling and investigate the potential mechanisms are needed.

## Limitations

This was a nonrandomized observational study from a single center, and patients who had no EF reassessment 3 months after revascularization were excluded. Therefore, as with any other observational studies, ours might be limited from selection biases. Echocardiographic evaluation is affected by intra- and inter-observer variability. To minimize this limitation, patients with echocardiographic assessment out of the center were excluded. 96.8% of EF measurements were reported by Simpson and only 3.2% of EF measurements were by Teicholz. As the Echocardiography operators in our hospital were all well trained, this variability might be minor. Large scale, multicenter and prospective studies are needed to confirm our finding.

## Conclusions

In current study, among CAD patients with reduced EF, approximately 45% of patients are likely to have LVESD reduction more than 7% after revascularization. A 7% reduction in LVESD had the best predictive accuracy for long-term survival. Combination of LV systolic function recovery and structure remodeling i.e., EF improvement and LVESD reduction, provided more precise risk stratification.

## Data Availability

The raw data supporting the conclusions of this article will be made available by the authors, without undue reservation.

## Acknowledgements

Thanks to all people contributing to the project of CRISIS.

## Abbreviations

AUC,: area under the curve
CABG: coronary artery bypass grafting
CAD: coronary artery disease
CRT: cardiac resynchronization therapy
EF: ejection fraction
HF: heart failure
HR: hazard ratio
LV: left ventricular
LVEDD: left ventricular end-diastolic dimension
LVESD: left ventricular end-systolic dimension
LVESV: left ventricular end-systolic volume
MI: myocardial infarction
PCI: percutaneous coronary intervention
ROC: Receiver-operating-characteristic

